# Synchrony-Division Neural Multiplexing: An Encoding Model

**DOI:** 10.1101/2021.10.29.21265658

**Authors:** Mohammad R. Rezaei, Milos R. Popovic, Steven A Prescott, Milad Lankarany

## Abstract

Cortical neurons receive mixed information from collective spiking activities of primary sensory neurons in response to a sensory stimulus. A recent study demonstrated that the time underlying the onset-offset of a tactile stimulus and its varying intensity can be respectively represented by synchronous and asynchronous spikes of S1 neurons in rats. This evidence capitalized on the ability of an ensemble of homogeneous neurons to multiplex, a coding strategy that was referred to as synchrony division multiplexing (SDM). Although neural multiplexing can be conceived by distinct functions of individual neurons in a heterogeneous neural ensemble, the extent to which nearly identical neurons in a homogeneous neural ensemble encode multiple features of a mixed stimulus remains unknown. Here, we present a computational framework to provide a system level understanding on how an ensemble of homogeneous neurons enable SDM. First, we simulate SDM with an ensemble of homogeneous conductance-based model neurons receiving a mixed stimulus comprising slow and fast features. Using feature estimation techniques, we show that both features of the stimulus can be inferred from the generated spikes. Second, we utilize linear nonlinear (LNL) cascade models and calculate temporal filters and static nonlinearities of differentially synchronized spikes. We demonstrate that these filters and nonlinearities are distinct for synchronous and asynchronous spikes. Finally, we develop an augmented LNL cascade model as an encoding model for the SDM by combining individual LNLs calculated for each type of spike. The augmented LNL model reveals that a homogeneous neural ensemble can perform two different functions, namely, temporal- and rate-coding, simultaneously.

## I. Introduction

Transmitting multiple signals over a single communication channel increases channel bandwidth and enhances coding efficiency [1, 2]. Similar to digital communication systems, the brain utilizes different forms of multiplexing – in different brain regions and in regard to different stimuli – to represent multiple features of a stimulus by a neural code [2]. For example, in the auditory sensory system, the frequency and intensity of a periodic stimulus are encoded by the phase-locked spikes and the probability of spiking per stimulus cycle, respectively[3]. Similarly, the frequency and intensity of vibrotactile stimuli are represented by the timing and rate of spikes in the somatosensory cortex [4]. Recently, differentially synchronized spiking neurons of the primary somatosensory cortex was shown to enable multiplexed coding of low- and high-contrast features of tactile stimuli [5]. Despite various forms of neural multiplexing, a thorough understanding of how the brain enables multiplexing remained undiscovered. Specifically, functional characteristics – in the sense of linear or nonlinear filtering properties – of a neural ensemble that multiplexes different features of a stimulus is yet to be uncovered. Different features of stimuli, like the intensity, frequency, onset and offset, etc., dictate which multiplexing strategies are most appropriate [4]. In addition to the properties of stimuli, heterogeneity of neurons in a population code enables different neurons to encode different stimulus features. The functional properties of a heterogeneous neural ensemble, which includes neurons with different functions, e.g., integrators vs. coincidence detectors, might be fully described by the dynamics of individual neurons. For example, an ensemble of heterogeneous cochlear nuclei in the auditory cortex is composed of two anatomically distinct sub-nuclei, namely, the magnocellular- and the angular-nucleus, each of which selectively encodes a specific feature of the stimulus. The magnocellular nucleus selectively encodes stimulus frequency with a temporal code by implementing high-pass filter whereas angular nucleus selectively encodes stimulus intensity with a rate code by implementing a low-pass filter [6]. In contrast to heterogeneous neural ensemble, functional characteristics of an ensemble of homogenous neurons, which includes neurons with nearly identical functions, cannot be identified based on the properties of individual neurons solely [4, 5]. For example, in synchrony division multiplexing (SDM) [5], information about slow and fast stimulus features were respectively represented by asynchronous and synchronous spikes of the same neurons. Thus, this form of multiplexing suggests that both slow and fast features of the stimulus can be encoded by homogeneous (identical) neurons that operate in a hybrid mode [5], i.e., neither low-pass nor high-pass filtering the stimulus [7, 8]. Thus, a challenging question is that whether multiplexing (like SDM) in a homogeneous neural ensemble reveals system-level functions beyond those performed by individual neurons [5].

In this paper, we utilize conductance-based and linear nonlinear (LNL) cascade models to establish a theoretical framework to address this question [9-12]. First, we use conductance-based models and construct a homogeneous neural ensemble that multiplexes slow and fast features of a common stimulus using asynchronous and synchronous spikes, respectively. Using the LNL cascade models, we explore whether different linear filters and static nonlinearities are associated with different types of spikes. We show that a low-pass filter followed by a nonlinearity with mild slope generates asynchronous spikes whereas a high-pass filter followed by a nonlinearity with steep slope detects fast features of the stimulus by generating synchronous spikes. Then, we develop an augmented LNL model for SDM by integrating LNL models underlying each type of spike.

## II. Results

In the present paper, we developed an augmented LNL cascade model as an encoding model for the SDM [5]. Conductance-based neuron models were used to create an ensemble of homogeneous neurons whose input (mixed stimulus) – output (spikes) relationship was estimated by the augmented LNL model.

As shown in **Figure 1A**, we construct an ensemble of homogenous neurons with 30 Morris-Lecar (ML) neuron models (see Methods) all of which receive a common mixed signal comprising slow and fast features as well as independent physiologically-realistic conductance noise [5, 13]. The parameters of the ML model were selected in a way that all neurons operate in a hybrid mode [14]. Spikes generated by an ensemble of ML neurons were used to fit the augmented LNL model in which two separate LNL models were combined to represent rate and temporal codes simultaneously. This study shows that an ensemble of homogeneous neurons utilizes different strategies to generate synchronous and asynchronous spikes which enable simultaneous coding of fast and slow features of a mixed stimulus, respectively. Although the biophysical mechanisms underlying implementation of SDM by an ensemble of homogeneous neurons is still unknown, the two-stream augmented LNL model provides a system-level understanding of the SDM function.

**Figure 1.**
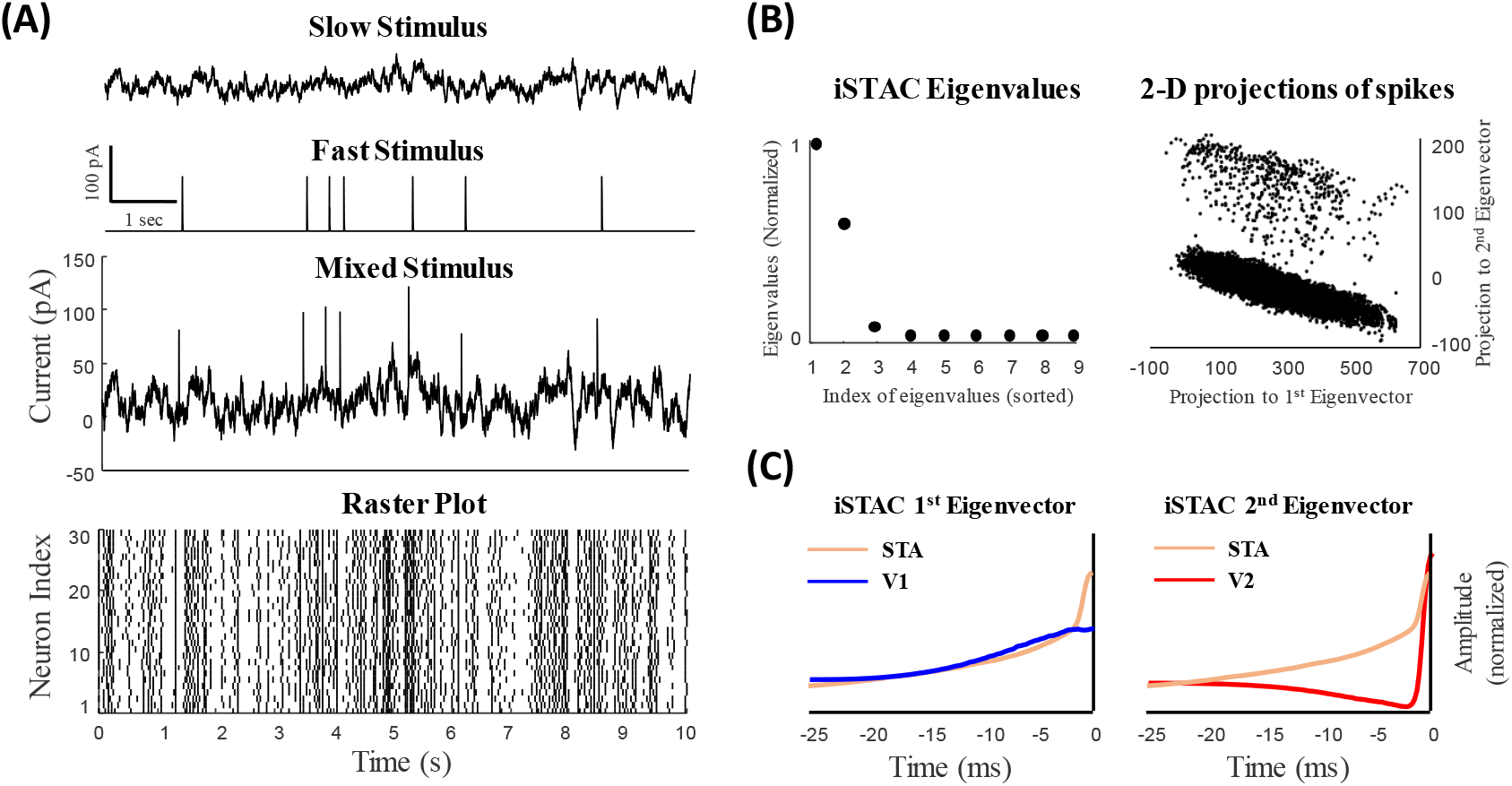
Slow and fast features of a mixed signal can be inferred from responses of a homogeneous ensemble of neurons using the iSTAC method. (**A**) Slow and fast signals constructing a mixed signal. (A, Bottom) Sample raster plot of 30 model neurons receiving the common mixed signal (and independent noise). Spikes evoked by the fast and slow signals cannot be distinguished visually. (**B**) The iSTAC method was applied to spike-triggered mixed signal and eigenvalues and eigenvectors were obtained (see Methods). (B, Left) The eigenvalues of the iSTAC matrix reveals two significant components of the population code. (B, Right) The projection of spike-triggered mixed signal onto the main eigenvectors of the iSTAC matrix. Two clusters can be visually distinguished. (**C**) The 1^st^ and 2^nd^ eigenvectors of the iSTAC matrix, *V1* and *V2*, respectively, are shown against the spike-triggered average (STA). *V1* resembles the STA filter reflecting slowly-varying changes in the signal. Unlike *V1, V2* resembles a high-pass filter (differentiator) which reflects fast features of the mixed signal.

### II. A. Different temporal filters map distinct features of a mixed stimulus

To explore how slow and fast features of the stimulus are encoded by spikes of an ensemble of neurons, we used well-known feature space estimators like the spike-triggered average (STA) [15, 16] or information-theoretic spike-triggered average and covariance (iSTAC) to reveal temporal characteristics of neurons in response to a stimulus[17].

The STA filter is a precise and unbiased predictor for a neural population given a stationary and single dimension stimulus [16]. However, it fails to provide precise predictions when the dimensionality of the stimulus is larger than one. For example, in retinal ganglion cells the STA cannot predict the neural response of both ON and OFF cells given a mixed input comprising more than one feature. To explore other possible subspace features of the neural response, we used the iSTAC method and calculated the optimal subspace features. The iSTAC quantifies the significance of subspaces based on the mutual information between stimulus and neural response [17]. In this method, we choose eigenvectors of the spike-triggered stimulus ensemble matrix more precisely by minimizing the Kullback-Leibler (KL) divergence between the eigenvectors of ensemble matrix and the raw stimulus distributions (see the method section for more details). In fact, the iSTAC maximizes information based on the first two moments of the spike-triggered stimulus ensemble and provides a unifying information-theoretic framework that captures the ensemble neuron activity in different subspaces. This provides an implicit model of the contribution of the nonlinear function mapping the feature space to the neural response. As shown in **Figure 1B (left)**, the iSTAC matrix calculated for the mixed stimulus has two significant eigenvalues whose underlying eigenvectors reveal two distinct temporal filters, namely, *v*_1_ and *v*_2_. The projection of the spike-triggered stimulus ensemble on *v*_1_ and *v*_2_, shown in **Figure 1B (right)**, reveals two distinct clusters.

In synchrony-division multiplexing, a mixed input signal containing slow and fast features drives an ensemble of neurons. The fast component of the stimulus whose neural representation is synchronous and sparse does not appear in the STA, as it averages out sparsely-occurring fast features of the stimulus [5]. However, unlike the fast signal, neural representation of the slow signal is asynchronous and dense, thus the STA filter mainly contains information of the slow components of the mixed signal [5]. Unlike the STA filter, the most informative subspaces selected by the iSTAC method behave as multi-space feature estimators and illustrate slow and fast features of the mixed stimulus.

**Figure 1C** shows that the STA filter calculated for the mixed stimulus mainly captures the slow feature of the signal, but cannot truly capture the dynamics of synchronous spikes. Unlike the STA filter, *v*_1_ and *v*_2_ of the iSTAC method illustrates slow and fast features of the mixed stimulus. As can be observed in **Figure 1C**, *v*_1_ is similar to the STA filter and represents the slow component of the stimulus, and *v*_2_ describes fast features of the stimulus (note that the STA filter was duplicated in **Figure 1C** (left and right) and compared with both *v*_1_ and *v*_2_).

### II. B. Low-dimension feature space of the neural response can be characterized by the STAs of synchronous and asynchronous spikes

Recently, it has been shown that synchronous and asynchronous spikes encode information of slow and fast features of a mixed stimulus (equivalent to that used in the present study), respectively [13, 18, 19]. Using an information theoretic approach, it was shown that synchronous and asynchronous spikes carry information in different time scales. By classifying spikes of a population of neurons into synchronous and asynchronous spikes, it was demonstrated that the STA filters underlying these spikes, namely, *μ*_*Async*_ and *μ*_*Sync*_, reflect the fast and slow features of the stimulus, respectively. **Figure 2A** shows classified synchronous (red) and asynchronous (blue) spikes in the raster plot.

**Figure 2.**
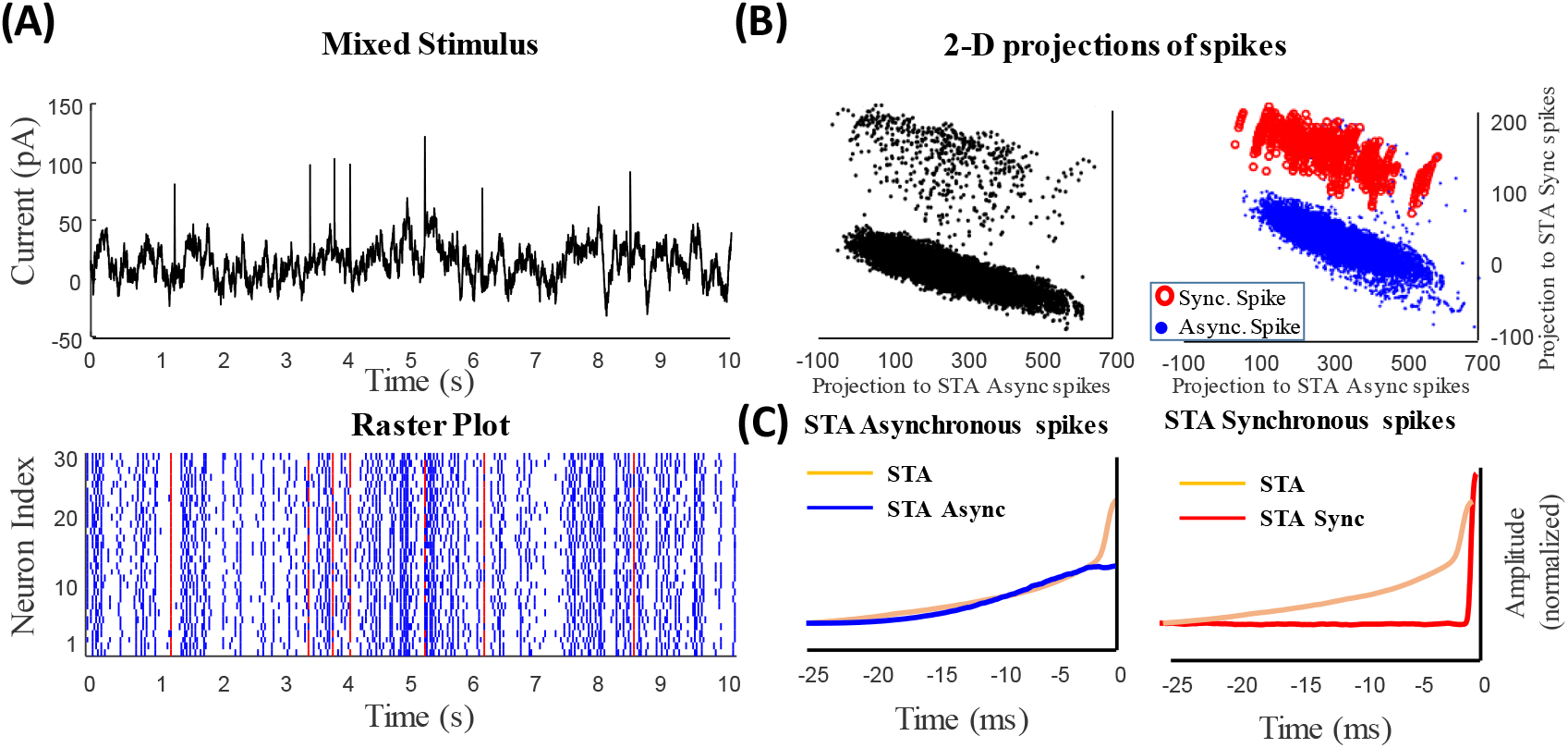
Synchronous and asynchronous spikes represent information of slow and fast features of the mixed signal, respectively. (**A**) Synchronous (red) and asynchronous (blue) spikes are distinguished by setting a threshold on the instantaneous firing rate calculated by a narrow kernel (see Methods). Synchronous spikes evoked by the fast signals can be distinguished visually. (**B**) The projection of spike-triggered mixed signal onto the STA_Sync_ and STA_Async_. Two (visually) distinguishable clusters belong to asynchronous spikes representing the slow feature of the signal (blue dots) and synchronous spikes representing the fast features (red circles). (**C**) The spike-triggered average of synchronous (red) and asynchronous (blue) spikes, namely, STA_Sync_ and STA_Async_, respectively, was shown against the STA of all spikes (similar to Figure 1. C).

Here, we compared these filters with those obtained by the iSTAC method. First, we tested if the projection of the spike-triggered stimulus ensemble on *μ*_*Async*_ and *μ*_*Sync*_ creates two distinct clusters similar to that projected on *v*_1_ and *v*_2_. As shown in **Figure 2B (left)**, two distinct and separable clusters were generated by *μ*_*Async*_ and *μ*_*Sync*_. More importantly, one can distinguish between these clusters by projecting synchronous- and asynchronous-spike-triggered stimulus ensemble on *μ*_*Async*_ and *μ*_*Sync*_. **Figure 2B (right)** reveals that these stimulus ensembles are separable and mutually exclusive. **Figure 2C** shows temporal patterns of *μ*_*Async*_ and *μ*_*Sync*_ versus the STA filter. As expected, *μ*_*Async*_ resembles the STA filter, indicating slow features of the stimulus, and *μ*_*Sync*_(similar to *v*_2_) describes abrupt changes in the stimulus.

Furthermore, to investigate the functional roles of the above filters, we tested how they contribute in the signal reconstruction. The reconstructed signal was obtained by the convolution of spikes - either all spikes for STA (**Fig 3A**) or *v*_1_ and *v*_2_ (**Fig 3B**) or asynchronous and synchronous spikes for *μ*_*Async*_ and *μ*_*Sync*_ respectively (**Fig 3C**). **Figure 3** illustrates 10 sec sample of the reconstructed signal using these methods. As clear in **Figure 3.B**, the signal reconstructed by *v*_1_ and *v*_2_(iSTAC method) resemble that generated by *μ*_*Async*_ and *μ*_*Sync*_, and both of these signals better capture fast features than that obtained by the STA filter, indicating the functional relevance between these filters.

**Figure 3.**
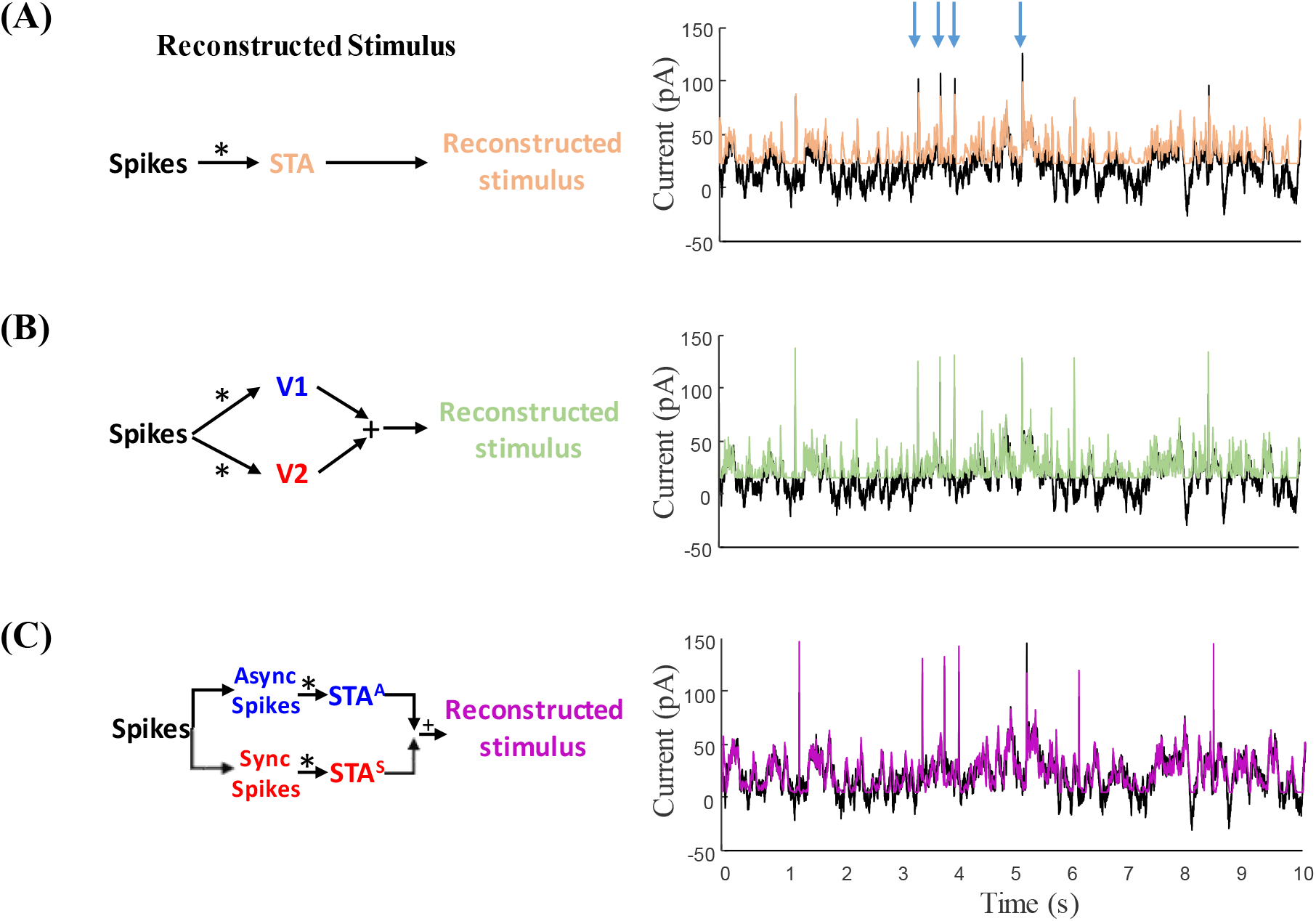
Block diagram of decoding the mixed signal from spikes by (**A**) the STA filter (light brown), (**B**) a weighted sum of the 1^st^ and 2^nd^ eigenvectors of the iSTAC method (light purple), and (**C**) a weighted sum of filtered asynchronous spikes (by STA _Async_) and filtered synchronous spikes (by STA_Sync_) (purple). Original mixed signal overlaid with black color in the plots. As can be seen in these figures, the reconstructed signal based on STA_Sync_ and STA_Async_ – similar to that obtained by eigenvectors of iSTAC method– can capture both slow and fast components of the signal accurately.

### II. C. Different nonlinear functions are associated with synchronous and asynchronous spikes

Given different temporal filters underlying synchronous and asynchronous spikes, we sought how these filters map fast and slow features of the mixed stimulus to the firing rate of an ensemble of conductance-based model neurons. Moreover, since the dynamics of a neural ensemble is not fully linear, these linear filters are not sufficient to project the stimulus to spikes. We utilized a well-known phenomenological model, namely, the LNL cascade model, which uses a linear stimulus filter followed by a static nonlinear transformation, to estimate the firing rate of an ensemble of neurons. **Figures 4** and **5** (panel A in both figures) show the LNL diagram for asynchronous and synchronous spikes, respectively. We tested if a pair of linear filter and static nonlinearity is different for synchronous and asynchronous spikes given a common mixed signal. We obtained static nonlinearity functions for synchronous and asynchronous spikes by applying *μ*_*Sync*_ and *μ*_*Asyncn*_ filters to the mixed stimulus (*s*) and mapping their outputs (through the nonlinearity) to the PSTHs of synchronous and asynchronous spikes, respectively:

**Figure 4.**
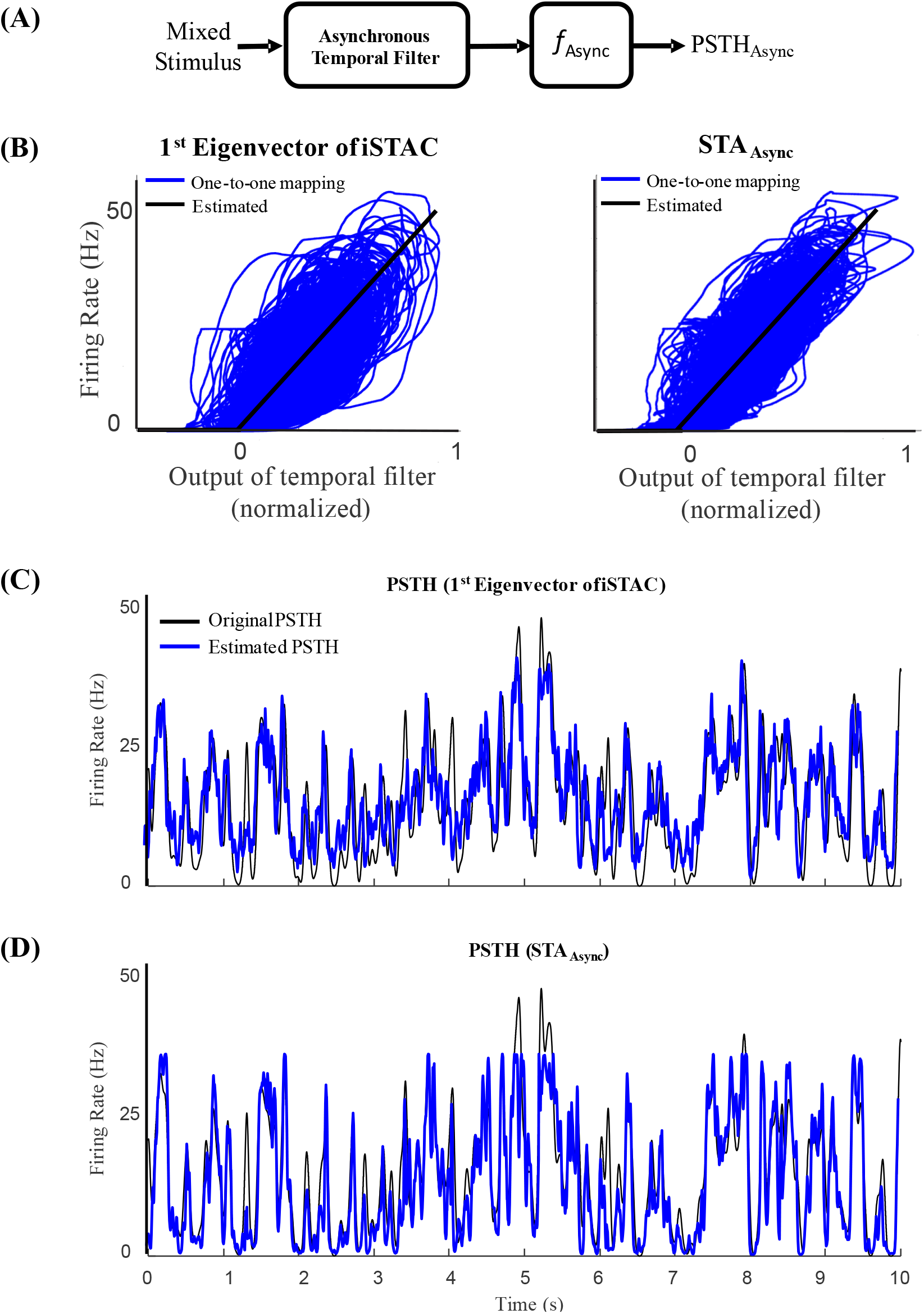
Static nonlinearities underlying asynchronous spikes. (**A**) Blog diagram of LNL model for asynchronous spikes. (**B**) Static nonlinearity calculated for the asynchronous spikes is obtained by mapping the output of filtered stimulus to the instantaneous firing rate of asynchronous spikes (calculated by a wide kernel, σ = 25 msec). Static nonlinearity calculated based on 1^st^ eigenvectors of the iSTAC method, *V1*, (Left) and STA_Async_ (Right). The solid black shows fitted rectifiers. (**C**) The PSTHs constructed by the fitted nonlinearities based on *V1* were drawn against the PSTH of asynchronous spikes. **(D)** The PSTHs constructed by the fitted nonlinearities based on STA_Async_ were drawn against the PSTH of asynchronous spikes.

**Figure 5.**
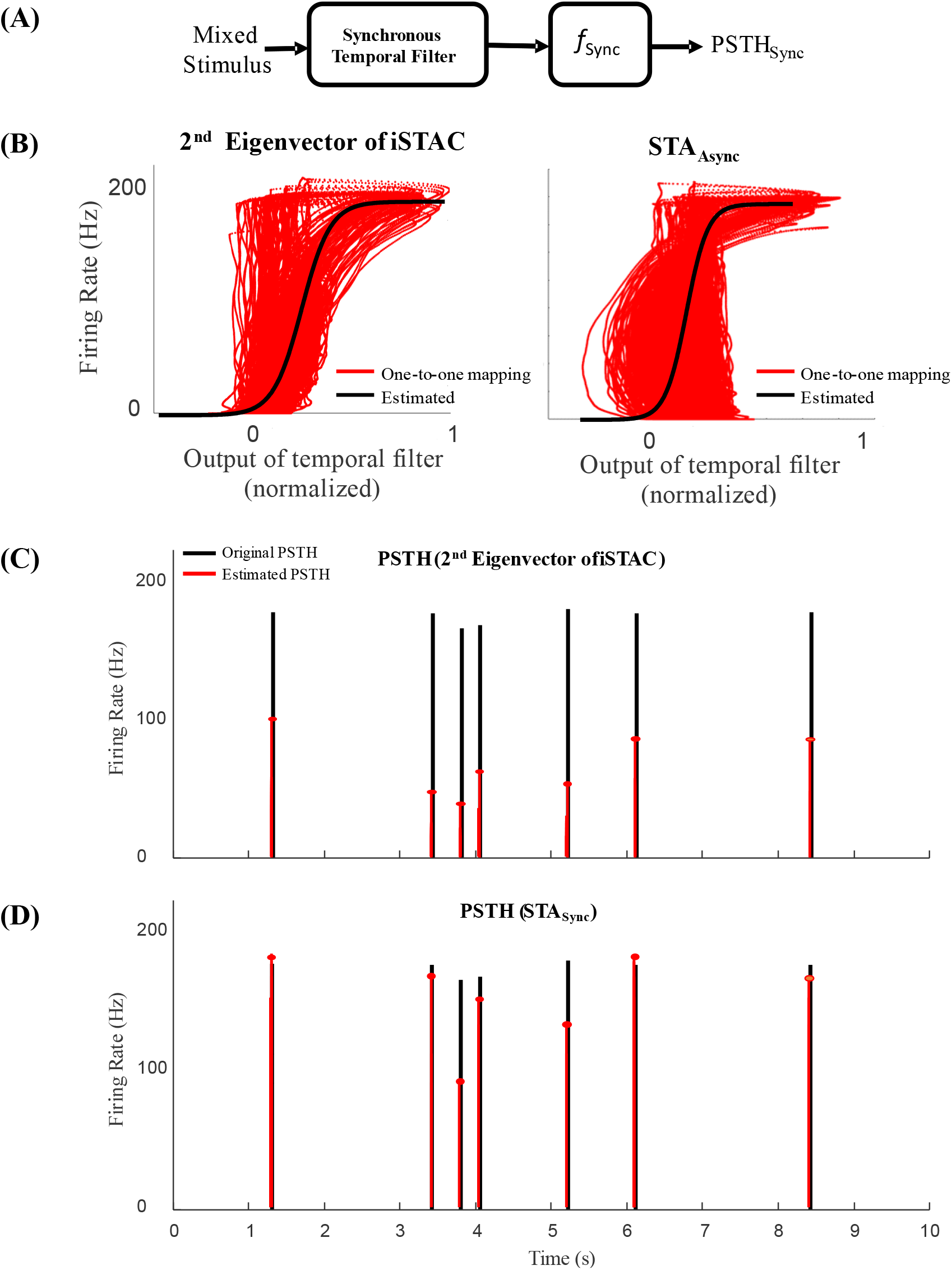
Static nonlinearities underlying synchronous spikes. (**A**) Blog diagram of LNL model for synchronous spikes. (**B**) Static nonlinearity calculated for the synchronous spikes is obtained by mapping the output of filtered stimulus to the instantaneous synchronous events (calculated by a narrow kernel, σ = 1 msec). Static nonlinearity calculated based on 2^nd^ eigenvectors of the iSTAC method, *V2*, (Left) and STA_Sync_ (Right). The solid black shows fitted sigmoid functions. (**C**) The PSTHs constructed by the fitted nonlinearities based on *V2* were drawn against the PSTH of synchronous spikes. **(D)** The PSTHs constructed by the fitted nonlinearities based on *V2* were drawn against the PSTH of synchronous spikes.

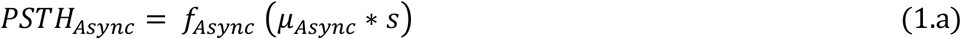

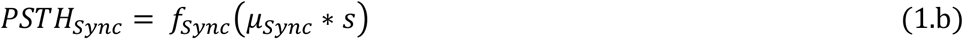

where *f*_*Async*_(*x*) and *f*_*Sync*_(*x*) are the nonlinearities associated with asynchronous and synchronous spikes, respectively.

**Figures 4(B)** and **5(B)** show respectively the raw nonlinearities for asynchronous and synchronous spikes that correspond to the mapping of every single point of the output of linear filters (x-axis) to the values of PSTHs (y-axis). For the nonlinearities underlying asynchronous and synchronous spikes, we fitted the ReLU nonlinearity and sigmoid functions, respectively [20]. The nonlinearly associated with asynchronous, *f*_*Async*_(*x*), has mild slope and broad dynamic range, enabling rate-modulated coding. In contrast, the nonlinearly underlying synchronous spikes, *f*_*Sync*_(*x*)), has steep slope and narrow dynamic range, enabling event (abrupt changes) detection. Although more sophisticated nonlinear functions could provide better fits, we chose simple and well-established nonlinear functions to highlight the difference in shapes of nonlinearities underlying rate-versus temporal-codes in the context of SDM. The instantaneous firing rates of each type of spike can be constructed by passing the output of temporal filter through the fitted nonlinearities. These firing rates were estimated and drawn against the PSTHs of asynchronous and synchronous spikes in **Figures 4(C-D) & 5(C-D)**, respectively. As shown in these figures, the nonlinear functions and estimated PSTHs underlying temporal filters obtained by the iSTAC (*V1* and *V2*) and classified spikes (*μ*_*Async*_ and *μ*_*Sync*_) are nearly identical.

### II. D. An augmented LNL cascade model for synchrony-division multiplexing

The LNL cascade models were utilized to encode specific features of a mixed stimulus by synchronous or asynchronous spikes. As shown in the previous sections, temporal filters and nonlinear transformations of either types of spikes were distinct and estimated by separate LNL cascade models. Here, we sought if a combination of these cascade models, i.e., an augmented LNL model, could accurately encode different features of a mixed stimulus through different types of spikes. We developed a two-stream LNL cascade model that combines the PSTHs of synchronous and asynchronous spikes to reconstruct the mixed PSTH of both type of spikes, as:

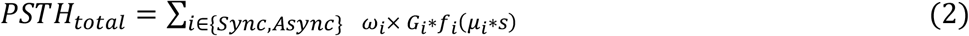

where *ω* is the combination weight for each stream of reconstructed PSTHs. To reduce the model complexity and promote smoothness in the output, we applied parameterized Gaussian kernels, *G*_*Async*_ = *Gaussian* (0, *σ*_*Async*_) and *G*_*Sync*_ = *Gaussian* (0, *σ*_*Sync*_), to the reconstructed PSTHs in each stream [21, 22]. The augmented LNL model simultaneously encodes slow and fast features of the stimulus by asynchronous and synchronous spikes, respectively. **Figure 6(A)** shows the block diagram of the augmented LNL model. This model implies that the low-pass filter and shallow non-linearity underlying asynchronous spikes are required to produce the rate code. In contrast, the high-pass filter and sigmoid nonlinearity of synchronous spikes are necessary to preserve reliable spike times underlying fast features of the stimulus. Taken together, the augmented LNL model makes the coexistence of the rate- and temporal-codes happen to encode multiple features of a mixed stimulus. To capitalize on the significance of temporal filters and nonlinearity transformations of each type of spike in estimating the total firing rate of a neural ensemble, we compared the performance of augmented LNL model with that of a conventional one stream LNL. **Figure 6(B-D)** shows the estimated firing rate of three methods, namely, Poisson GLM and augmented LNL models (**Figure 6(C-D)**) (used Section II.C), against the PSTH of ensemble of neurons. As can be observed, the firing rate estimated by the augmented LNL models can better capture both the rate of asynchronous spikes and timing of synchronous events compared to those estimated by one stream Poisson GLM.

**Figure 6.**
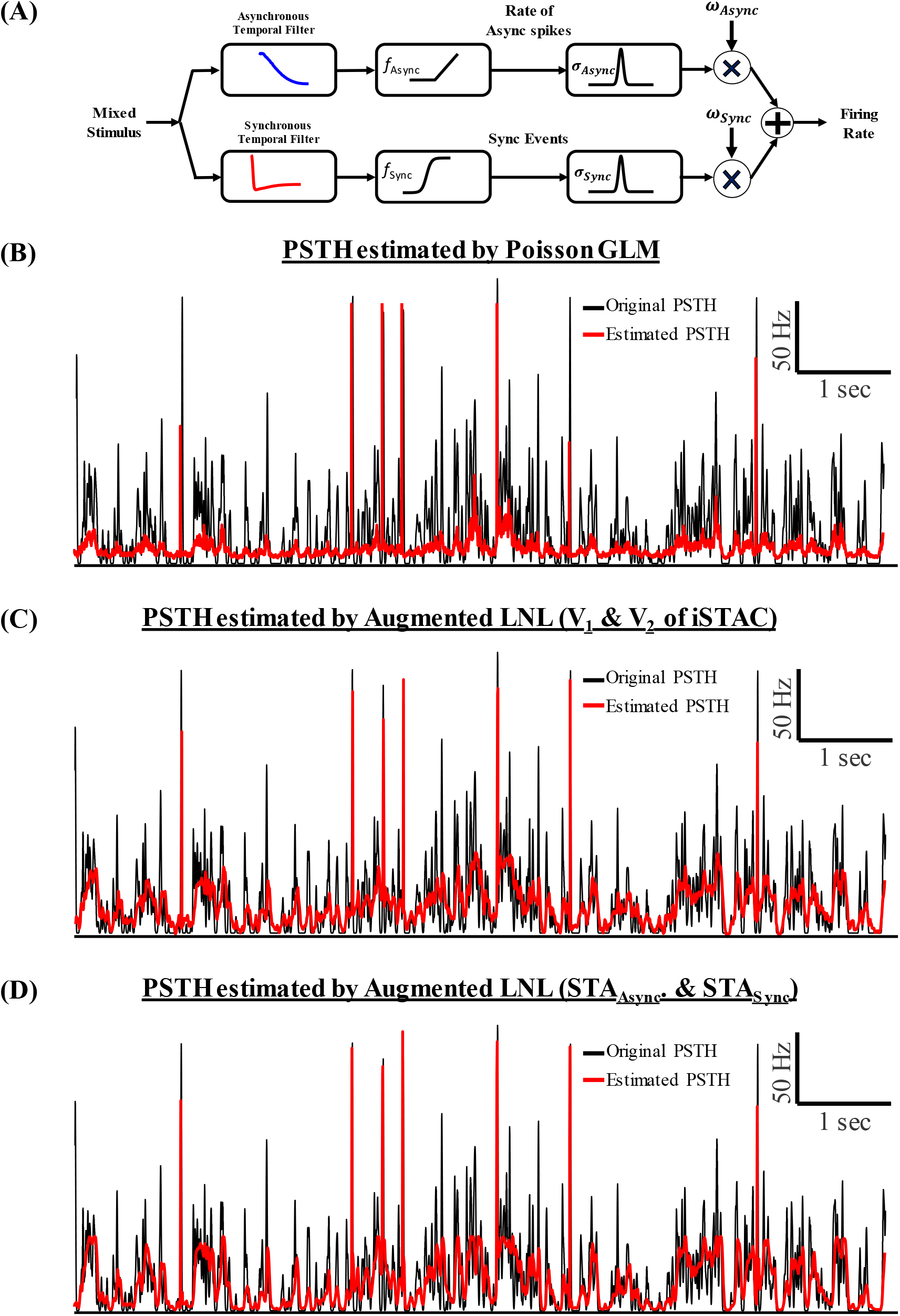
Two stream LNL model, referred to as augmented LNL model, enables co-existence of temporal- and rate-codes. (**A**) Blog diagram of the augmented LNL model for combining rate of asynchronous spikes and events of synchronous spikes. (**B**) The PSTHs estimated by a conventional Poisson GLM (red) were shown against the original PSTH (calculated by 1 msec Gaussian kernel). **(C)** The PSTHs estimated by the segmented LNL using temporal filters of iSTAC method. **(D)** The PSTHs estimated by LNL using the segmented LNL using STA_Async_ and STA_Sync_.

## III. Discussion

The ability of an ensemble of homogeneous cortical neurons to multiplex multiple features of a mixed stimulus was studied in [5]. However, specific encoding functions underlying these neurons were not determined. In this paper, we presented a computational framework to provide a system level understanding of the encoding mechanism underlying SDM. We used conductance-based neuron models to construct a homogenous neural ensemble that encodes slow and fast features of a mixed stimulus through asynchronous and synchronous spikes, respectively. To elucidate the contribution of slow and fast features of the mixed stimulus on the spikes generated by model neurons, we calculated most significant subspaces (eigenvectors) of spike-triggered stimulus matrix using the iSTAC method. We demonstrated that the calculated first and second eigenvectors resemble slow and fast features of the stimulus, respectively. Furthermore, the projection of spike-triggered stimulus matrix on these eigenvectors created two distinct clusters. We tested if these clusters can be characterized by synchronous and asynchronous spikes. By computing the spike-triggered average (STA) filters of synchronous and asynchronous spikes, and projecting this matrix on these filters, we clearly separated those clusters. Furthermore, we fitted a LNL model to each type of spikes. Similar to distinct temporal filters for synchronous and asynchronous spikes, their static nonlinearities have different properties. We found that the nonlinearity associated with asynchronous spikes is very shallow and can be approximated by a linear function. On the other hand, the nonlinearity associated with synchronous spikes has a very steep slope and can be approximated by highly nonlinear functions like sigmoid function. Finally, we developed an augmented LNL model to capture both dynamical characteristics of synchronous and asynchronous spikes and reconstruct the PSTH of all spikes.

### Subspace Feature Estimators; iSTAC vs. STC

To explore more than one subspace features for stimulation-evoked neural responses, we compared the performance of the STC and iSTAC methods. One can find most informative subspaces that maximize the mutual information between stimulus and response [23, 24]. Nevertheless, an accurate estimation of mutual information requires a large amount of data although no guarantee for optimal estimation can be necessarily expected[24]. A conventional way to find these subspaces is to calculate those related to the most significant eigenvectors of the spike-triggered covariance (STC) matrix [16]. The eigenvectors of the STC matrix provide analytic expressions for filter estimation using the moments of the stimulus and spike-triggered stimulus distribution [16, 17]. However, this method does not incorporate joint information between the mean and the variance, and also there is no specific measurement for selecting the most significant subspaces based on that information. We calculated the most important eigenvectors of the STC matrix underlying the mixed stimulus and neural response (see **II.B**, details are provided in **Methods**). The first eigenvector of the STC matrix was similar to the STA of all spikes (see **Figure 7**) including both slow and fast features of the stimulus. Unlike the first eigenvector of the STC matrix, the second eigenvector comprised fast features of the stimulus. As shown in Figure **7**, the 2-D projection of the spike-triggered stimulus matrix on eigenvectors of the STC matrix cannot be clearly separated in two distinct clusters.

**Figure 7.**
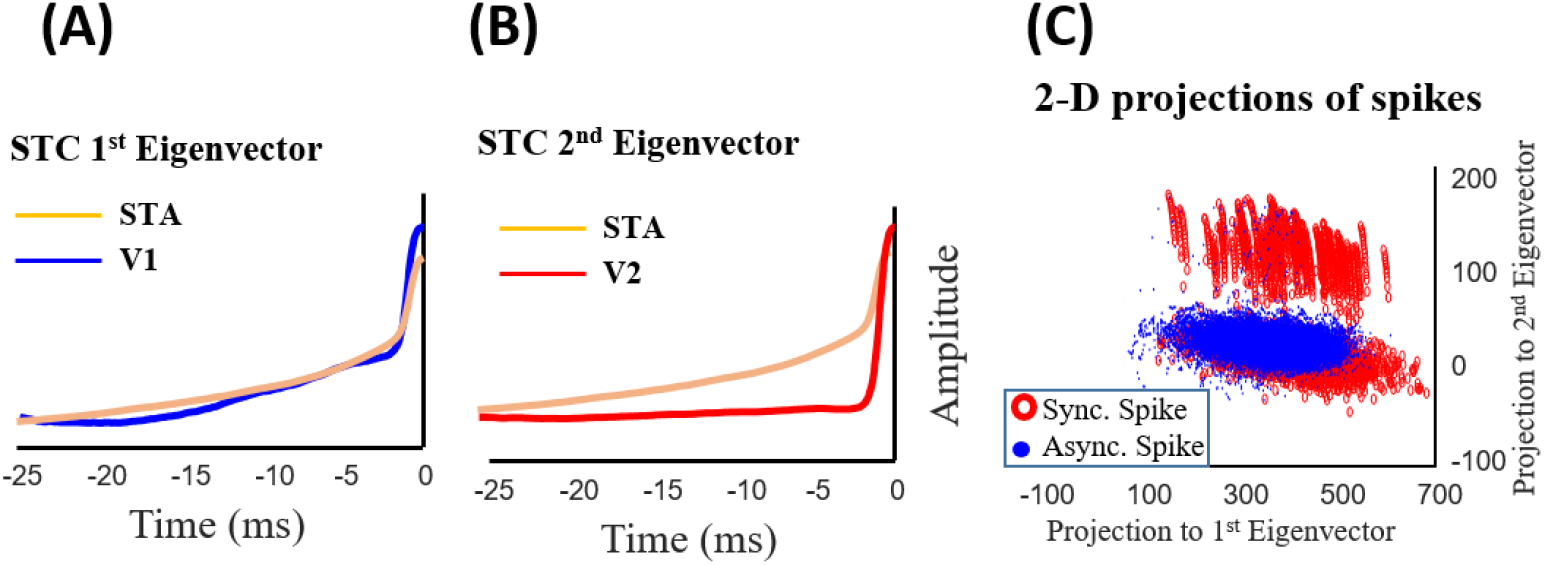
Slow and fast features decomposition of a mixed signal of a homogeneous ensemble of neurons using the STC method. (**A**) The projection of spike-triggered mixed signal onto the 1^st^ eigenvector and (**B**) 2^nd^ eigenvector of the STC matrix. (**C**) The 1^st^ and 2^nd^ eigenvectors of the STC matrix, *V1* and *V2*, respectively, are shown against the spike-triggered average (STA) calculated using all spikes.

To avoid this problem, we used the iSTAC method that allows to choose eigenvectors of the spike-triggered stimulus ensemble matrix more precisely by minimizing the Kullback-Leibler (KL) divergence between the eigenvectors of this matrix and that obtained by raw stimulus distributions [17]. It is to be noted that the whitening transformation is usually used before finding subspaces of the spike response. One can use whitening transformation to calculate uncorrelated and normalized subspaces (for both STC and iSTAC methods). However, due to the type of the mixed stimulus (i.e., structured and not a random process), we found that eliminating this transformation results in more representative subspace features as shown in **Figure 1**. We compared the 2-D projection of the spike-triggered stimulus matrix on eigenvectors of the iSTAC method with and without whitening. It can be clearly observed that the iSTAC without whitening can better separate the 2-D space.

### Choice of Static Nonlinearity in the LNL Model

The static nonlinearities obtained in the augmented LNL model can describe why synchronous and asynchronous spikes are associated with different functions. For example, the smoothness and linear behavior of *f*_*Async*_(*x*), for *x>0*, generates a smooth PSTH for asynchronous spikes which linearly encodes to the intensity of the stimulus. In contrast, the sigmoid-like nonlinearity of synchronous spikes, *f*_*Sync*_(*x*), maintains the PSTH of synchronous spikes very sparse which nonlinearly detects abrupt changes of stimulus. It is worth mentioning that more flexible nonlinear functions could provide better fits for representing the PSTH of synchronous and asynchronous spikes. Of note, one can use deep neural networks (DNNs) to give more flexibility to the models. A DNN is simply a high-dimensional non-linear function estimator which gives a multilayer nonlinear function in the form of a neural network [25, 26]. To use that in our augmented LNL model or any LNL model, one can easily replace the nonlinearity estimator with a DNN.

### Generalized Linear Model (GLM) for Augmented LNL

The proposed augmented LNL can also be interpreted in the GLM framework. From this point of view, synchronous and asynchronous PSTHs are modeled by two separated GLMs with different random processes, which eventually combine their PSTHs linearly. The first GLM, filters the mixed stimulus with the first eigenvector of iSTAC and then by passing it through a nonlinearity and then a Gaussian random process (with a linear nonlinearity as its link function), it models the PSTH related to asynchronous spikes. Likewise, the second GLM, filters the mixed stimulus with the second eigenvector of iSTAC and then by passing it through a nonlinearity and then a Bernoulli random process (with a sigmoid nonlinearity as its link function), it models the PSTH related to synchronous spikes (See the method section for more details about GLMs). Or, we even simply can interpret the augmented LNL as a single Poisson GLM, with two input filters (the first two eigenvectors of iSTAC) and a Poisson random process at the end (see the appendix A for more details). To reach the optimal parameters set for the model and avoid computational complexity, we use parametric models for the static nonlinearities [9]. We also can use more flexible parametric function (with parameter set θ) like ex-quadratic function *f*_*θ*_(*x*) as the static nonlinearities. By using ex-quadratic function as nonlinearities we eventually need to optimize a convex cost function, Which gives the optimum parameters set θ for the nonlinearity and can be optimally optimized by a maximum-likelihood (ML) algorithm (details in the Appendix B) [9, 27, 28].

## Materials and Methods

### Simulated mixed input

According to the feasibility of neural systems to multiplexed coding, we simulated the activity of a homogeneous neural ensemble in response to a mixed-stimulus to explore how much information can be encoded by different patterns of spikes. Each neuron received a mixed signal (*I*_*mixed*_) which consists of a fast signal (*I*_*fast*_) and a slow signal (*I*_*slow*_). *I*_*fast*_ Stands for the timing of fast events or abrupt changes in the stimulus and was generated by convolving a randomly (Poisson) distributed Dirac-delta function with a synaptic waveform (normalized to the peak amplitude), *τ*_*rise*_ = 0.5 *ms*, and *τ*_*fall*_ = 3 *ms*. Fast events occurred at a rate of *∼* 1 *Hz* and were scaled by *a*_*fast*_ = 85 *pA*.

*I*_*slow*_ was generated by an OU process as follows.

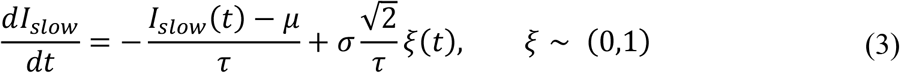

where *ξ* is a random number drawn from a Gaussian distribution, *τ* = 100 *ms* is the time constant of the slow signal that produces a slow-varying random walk with an average of μ = 15 *pA* and a standard deviation of *σ* = 60 *pA*. The mixed signal (*I*_*mixed*_) was obtained by adding *I*_*fast*_ and *I*_*slow*_, were generated independently. An independent noise (equivalent to the background synaptic activity) was added to each neuron, thus each neuron receives a mixed signal plus noise. Similar to, the noise (*I*_*noise*_) was generated by an OU process of *τ* = 5 *ms*, μ = 0 *pA*, and *σ* = 10 *pA*.

### Simulated neural ensemble and its response to mixed input

The neural ensemble consists of 30 neurons, each of them was modeled by Morris-Lecar equations [13, 29]. The equations of a single model neuron receiving a mixed-signal plus noise can be written as follows.

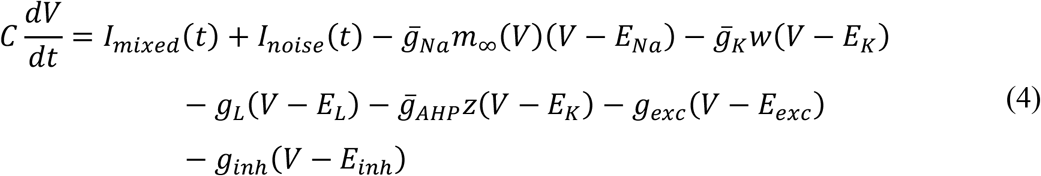

where,

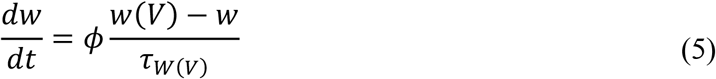

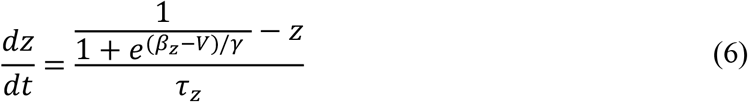

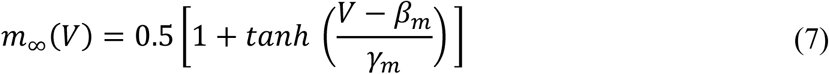

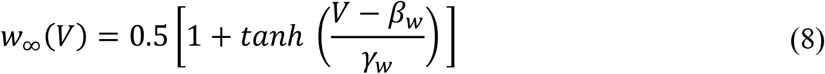

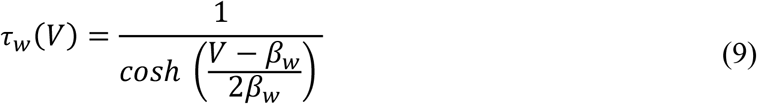

Where 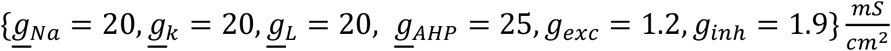, *β*_*m*_ = *−*1.2, *γ*_*m*_ = 18, *β*_*w*_ = − 19, *γ*_*w*_ *= 10 β*_*Z*_ = 0, *γ*_*Z*_ = 2, *τ*_*a*_ *=* 20 *ms,ϕ =* 0.15, *and* 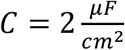. These parameters were set to ensure a neuron operates in a hybrid mode [30], i.e., an operating mode between integration and coincidence detection [5, 31]. The inclusion of background excitatory and inhibitory synaptic conductance in (2) reproduced a “balanced” high conductance state. The surface area of the neuron was set to 200 μ*m*^2^ so that *I*_*mixed*_ is reported in *pA*, rather than as a density[32, 33]. **Figure 1.A** Shows the mixed stimulus and the spiking activates of the ensemble of neurons in response to this stimulus.

### Generalized Linear Model (GLM) details

GLM model is a generalization of traditional linear models, which gives the neural encoding models more flexibility to capture nonlinear dynamics of neural activity. GLM contains three stages. The first stage is a linear mapping which consists of a set of *d* linear-filters, let’s assume ***K*** = [*k*_1_, …, *k*_*D*_], that maps high dimensional sensory stimulus *s* ∈ *R*^*M*^ into a low dimensional stimulus feature map *x* ∈ *R*^*D*^:

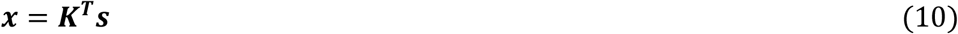

The second stage is a pointwise nonlinearity, *f*: *R*^*D*^ → *R*, which maps the linear features of *d* dimensions into a nonnegative spike rate:

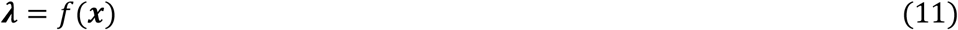

In the final stage, the number of spikes generated by a random process:

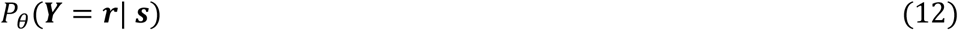

Where *Y* is random variable related to spikes occurrence, ***r*** is instantaneous firing rate, and the *θ*is parameter set of the random process

In simple words, by using GLM we approximate the instantaneous firing rate by considering feature from *D* dimensions instead of *M* dimensions:

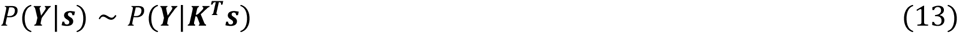

So, there are two set of parameters, the estimators (***K***) and the pointwise nonlinearity (*f*), which can be optimized to reach the desired model.

### STA and STC estimator

If we assume that *p*(*s*) is has zero mean, then the STA can be defined as the average of the stimulus given the instantaneous firing rate:

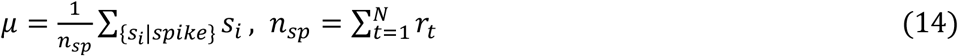

Where *N* is the total number of time points. The STA is an unbiased, consistent estimation which gives the direction in the stimulus space along which the means of *P*(***s***|*spike*) and *P*(***s***) differ most. The problem is The STA gives a single direction in stimulus space and leads to a single estimator which is not efficient to capture all information in the stimulus space (we previously discussed we have a mixed stimulus in this research). To involve other possible directions with maximally differences in variances between *P*(***s***|*spike*) and *P*(***s***) we can use eigenvectors of the STC matrix, defined as:

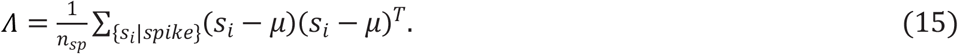

The STA and eigenvectors of the STC matrix can provide a basis, *K*, for a reduced dimensional model of the neural response.

### iSTAC estimator

There are two major problems with STA/STC, when we consider more than one direction for stimulus space. The first one is that STA is not orthogonal to STC eigenvectors and this increases the risk of losing information, and the second one is that the measure we use to select eigenvectors of STC is based on eigenvalues, which does not truly represent most informative directions. As we mentioned before the objective in iSTAC is to reduce KL divergence between Gaussian approximations to the spike-triggered and raw stimulus distributions. Therefore, we define *Q* as a Gaussian approximation of *P*(***s***|*spike*) based on the information contained only in the mean and covariance of the *P* as:

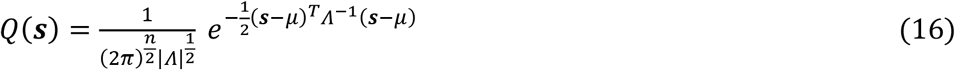

Where n is dimensionality of stimulus space. Now we drive KL divergence between *Q* and *P* as:

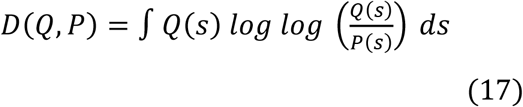

By considering that mean of *P* and *P. Q* is zero and have identity covariance (if not, we can use “whitening” technique) we can rewrite *D* in a simpler form as:

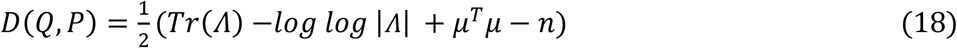

Where *Tr*(.) And |. | indicate trace and determinant, respectively.

Based on the fact that we are interested in *d* subspaces we can approximate the *D* with:

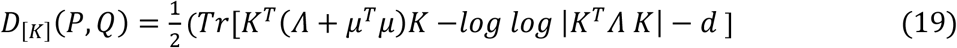

where *d* is the dimension of the interested subspaces.

So, in terms of finding the *d* most informative subspaces decomposed by STA and eigenvectors of STC we need to find *D*_[*K*]_(*P, Q*) for all subspaces and select the first *d* ones.

An important advantage of the iSTAC approach over traditional STA/STC analysis is that it makes statistically efficient use of changes in both mean and covariance of the response spaces.

## Data Availability

All data produced in the present study are available upon reasonable request to the authors

## Appendix A: The augmented LNL as a Poisson GLM

We assume the number of spikes in time are discrete events. By dividing the time horizon of the experiment, (0, *T*], into *K* (*K* is a large number) subintervals 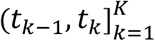 and consider *N*_*k*_ as number of the events in the time interval (*t*_*k*−1_, *t*_*k*_]. We can model the spike observation process with a point process by inhomogeneous Poisson distribution and parameter *λ*_*k*_ as:

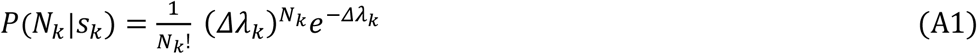

Where ⊿ is the width of the time-bins. By assuming that number of spikes are conditionally independents in time, we can write the whole observation process as

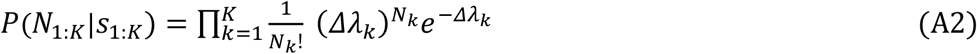

In other hand, we can model the *λ*_*k*_ in a way that captures effects of two features of the stimulus as linear combination of their effects in *λ*_*k*_ (discussed in the Result section), as

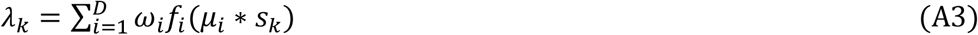

where, *θ* = {*ω*_1_, …, *ω*_*D*_} is our parameters set and *D* is the dimension of the feature map. Finally by using maximum-likelihood estimation we can tune the model parameters

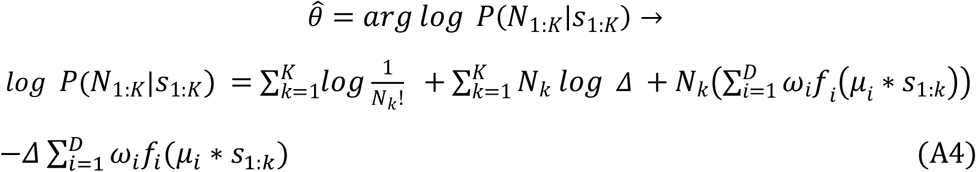

Based on Jensen’s inequality and considering that *log log x* is a concave function, we have:

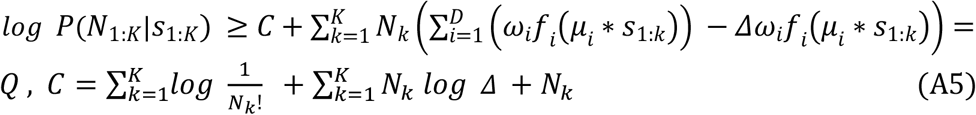

where *Q* is a lower bound for the Log-likelihood. So, we can find the *θ*s by maximizing the *Q* over them as

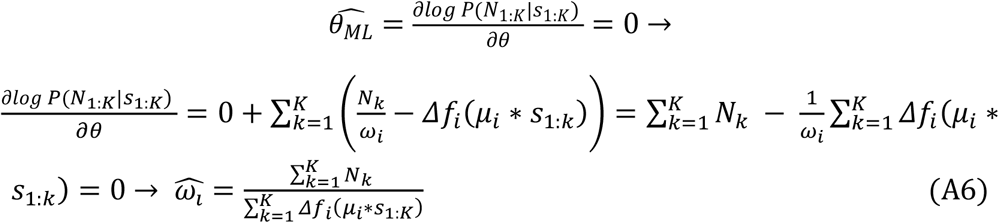

By estimating the model parameters, we reach a model for encoding the firing rate of the multiplexed spikes.

## Appendix B: Modeling nonlinearity (f(x)) by ex-quadratic functions

The estimators do dimensionality reduction task and map high dimensional sensory stimulus to a lower dimensional linear feature map = *K*^*T*^*s, K* is the basis of the feature map space. Based on the definition of GLM, mentioned above, we still need to find optimum model for *f*(*x*), *f*: *R*^*d*^ → *R*. By considering motivation in [4], a reasonable way is using exponential general quadratic function:

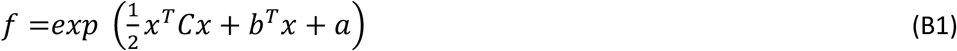

where *C* is a symmetric matrix, *b* is a vector, and *a* is a scalar. So, now we can use maximum-likelihood to optimize the parameters set, {*C, b, a*}. To do that we need to maximize the log-likelihood of observing spike given all spikes and the parameters set (*L* =*log log P*(*r*_1:*N*_|*s*_1:*N*_, *C, b, a*)). By assuming that spikes firing in time are independent then we can rewrite it as:

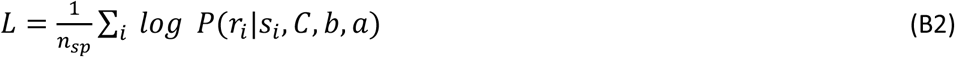

where *n*_*sp*_ is total number of spikes, so our objective is to maximize *L* by finding best parameters set:

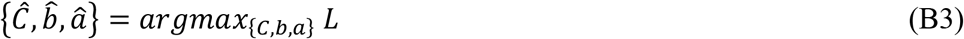

By following the optimization steps in [5] and assuming that the stimulus are drawn from *x* ∼ *N* (0, *ϕ*); the maximum-likelihood estimation of the parameters are:

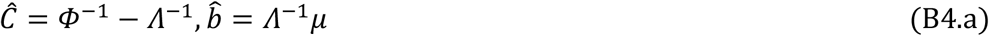

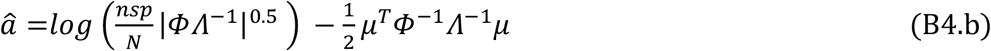

## Notes

### Competing Interest Statement

The authors have declared no competing interest.

### Funding Statement

This study did not receive any funding

